# Beware (surprisingly common) left-right flips in your MRI data: an efficient and robust method to check MRI dataset consistency using AFNI

**DOI:** 10.1101/19009787

**Authors:** Daniel R. Glen, Paul A. Taylor, Bradley R. Buchsbaum, Robert W. Cox, Richard C. Reynolds

**Affiliations:** Scientific and Statistical Computing Core, NIMH/NIH/DHHS, Bethesda, Maryland, USA; Rotman Research Institute at Baycrest, Toronto, Ontario, Canada; Department of Psychology, University of Toronto, Toronto, Ontario, Canada

## Abstract

Knowing the difference between left and right is generally assumed throughout the brain MRI research community. However, we note widespread occurrences of left-right orientation errors in MRI open database repositories where volumes have contained systematic left-right flips between subject EPIs and anatomicals, due to having incorrect or missing file header information. Here we present a simple method in AFNI for determining the consistency of left and right within a pair of acquired volumes for a particular subject; the presence of EPI-anatomical inconsistency, for example, is a sign that dataset header information likely requires correction. The method contains both a quantitative evaluation as well as a visualizable verification. We test the functionality using publicly available datasets. Left-right flipping is not immediately obvious in most cases, so we also present visualization methods for looking at this problem (and other potential problems), using examples from both FMRI and DTI datasets.

## INTRODUCTION

As part of the NIFTI dataset standard (Cox et al., 2004), orientation and location information were included in the file headers, to be able to reduce uncertainty in interpretation across software and systems. However, it is possible for mistakes to occur while recording information at the scanner, while interpreting DICOM fields, when converting to NIFTI or another format, or during a subsequent processing step. This can lead to seriously erroneous results (or to catastrophic results in clinical surgical cases).

While some header mistakes can be easily spotted visually (e.g., having incorrect voxel dimensions recorded), there are more subtle changes, particularly involving the positioning of the data within the acquisition field of view (FOV). The header contains the “orientation” parameters for how the data matrix is stored on disk, so that the rows and slices appear in the correct locations and view planes; an incorrect value can lead to axes that are switched (e.g., axial slices are interpreted as coronal ones) or flipped (e.g., the anterior part of the brain is labeled as posterior). Some of these problems can be recognized instantaneously, such as a flip in the anterior-posterior or inferior-superior axes in a whole brain acquisition; but others are much more subtle, such as left-right flips, due to the large-scale structural symmetry of the brain. Consider Fig. 1, which shows an axial slice of a T1w anatomical volume with both the original version of that subject’s EPI and a left-right flipped version (created by altering the orientation field of the file header). The correct orientation is often not visually obvious even when looking for this problem.

**Figure 1.**
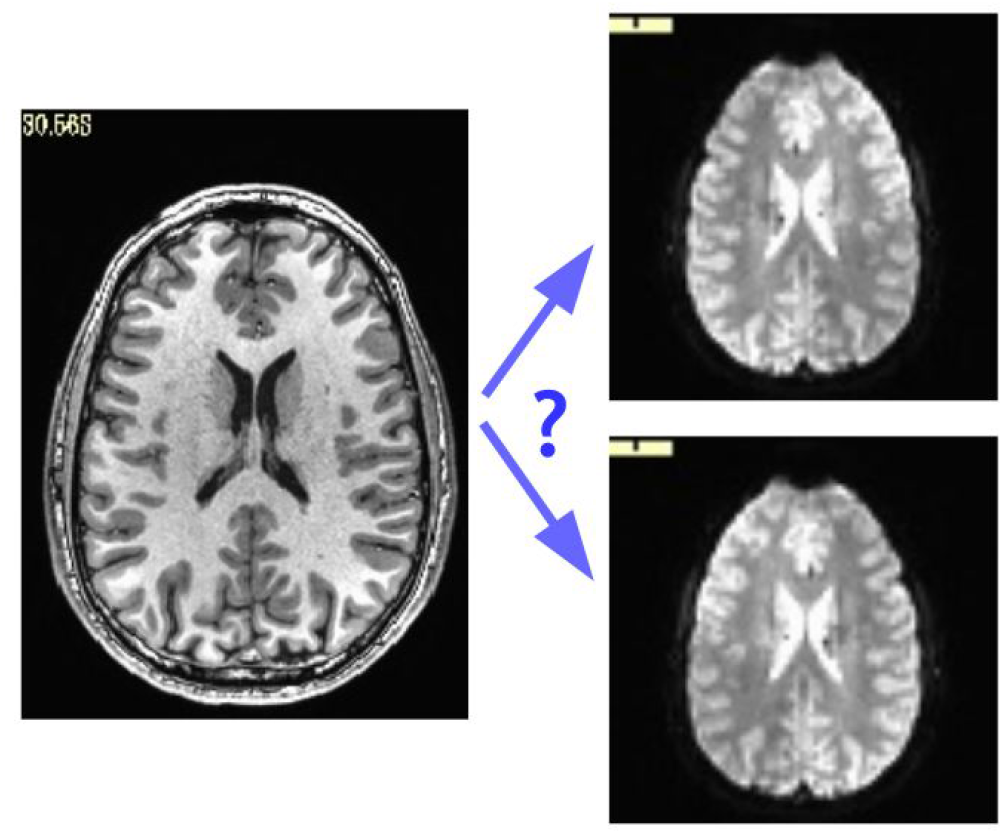
Axial slice of a T1w anatomical volume (left) with two versions of the same subject’s EPI volume (right): one is the original, and one is left-right flipped (through changing the orientation value stored in the file header). It is not immediately apparent which EPI image matches correctly with the anatomical, which is a problem for trusting results of analyses.

Here, we propose a simple method to detect relative left-right flips within pairs of MRI datasets (though it may also be more broadly applicable), available within the AFNI software package (Cox, 1996). The method applies to datasets with multiple acquisition types, such as EPI and T1w datasets acquired together for FMRI protocols, or DWI and T2w datasets for DTI protocols. How do we know that this problem actually occurs in practice? By using our approach, we have found systematic left-right flips in datasets submitted to each large public repository in which we have looked: the Functional Connectome Project (FCP) (Biswal et al., 2010), OpenFMRI (Poldrack et al. 2013), and ABIDE (Di Martino et al., 2014). These problems were verified by each notified consortium and subsequently fixed.^1^ We note this not to criticize these projects (indeed, the openness of these databases has made it possible to address these issues), but instead to point out how important it is for researchers to be able to check the basic properties of their data. This problem also exists in clinical imaging, with several studies reporting on rates of “laterality errors” in radiology (e.g., Bernstein et al., 2003; Sangwaiya et al., 2009; Landau et al., 2015; Digumarthy et al., 2018). If this fundamental problem can occur within large, public dataset collections that have been looked at by many people, as well as in clinical settings that affect patient outcome directly, then surely this is an issue that *all* researchers and neuroimagers should be aware of.

We present a simple diagnosis for this data issue. This efficient and robust check is available within the AFNI software package, along with several ways to visually verify the quantitative results. The visualization methods are shown using AFNI, but some of these methods are likely available in other software packages. In the Discussion, we note some of the potential causes of the flip issue.

## METHODS

The primary method introduced here is to check for left-right flipping through volumetric alignment: comparing the alignment cost function value between a pair of original datasets with the cost function of alignment when one dataset has been intentionally left-right flipped. This relies on having an appropriate cost function for the alignment. For instance, in FMRI studies, one generally wants to align a subject’s T1w anatomical with an EPI volume, which has a very different contrast pattern; T1w volumes in adult human datasets show decreasing brightness from white matter (WM) to gray matter (GM) and then to cerebrospinal fluid (CSF); in an adult human T2w EPI, the intensities are reordered relative to the T1w volumes, with CSF the brightest, then GM, and then WM. The reversal of the CSF relative intensities (i.e., the CSF is bright in EPI data yet dark in T1w datasets) is a key property that can be used for the advantage of alignment, in conjunction with aligning sulcal and gyral features. For these cases, AFNI uses the “local Pearson correlation (LPC)” cost function (Saad et al., 2009) or a variant “LPC+ZZ”, which have been shown to be robust in such cases. The LPC cost function is the negative of the sum of correlations computed over local regions (“patches”). Alignment then proceeds by optimizing for the minimal cost (negative correlation). The related “LPC+ZZ” cost function operates similarly but uses a combination of cost functions for its initial estimate and then finalizes the alignment parameters with the standard LPC cost^2^; this refinement has shown improvement in stability when the volumes being aligned have greater initial differences (e.g., large relative rotation). For aligning brains with similar tissue contrast (e.g., T2w anatomical with the b=0 s/mm^2^ volume in DWI datasets), AFNI typically uses the similarly robust “local Pearson absolute (LPA)” or related “LPA+ZZ” cost function, both using the absolute value of the local Pearson correlation.

The inputs for the check are simply the EPI and either anatomical volume; because the check is for relative flips, neither dataset is tagged as “correct” a priori. AFNI’s *align_epi_anat*.*py* program performs linear affine (12 degrees of freedom, by default) alignment between the volumes.^3^ There is no requirement for the datasets to be in particular coordinate or orientation systems; typically, they are in native/acquired coordinates. One simply provides an option flag “-check_flip”, and the program will generate cost function results and aligned volumetric datasets for the two cases: for the original volumes, and with one volume (e.g., the anatomical) left-right flipped. If the alignment is improved (lower cost function metric) for the intentionally left-right flipped data, then the flipped dataset is likely better, and an error is probably present in one of the original volume headers (NB: this method detects inconsistencies between volumes in a dataset, with further follow-up needed to determine which file is incorrect; see the Discussion for more on this). The cost function results and an ensuing recommendation about whether a dataset needs to be flipped (“NO_FLIP” or “DO_FLIP”), are saved in a text file. The aligned volume from each case is also output, for visual comparison and verification.

The left-right flip check can also be performed conveniently as part of the FMRI processing pipeline constructed by AFNI’s *afni_proc*.*py*. The option “-align_opts_aea -check_flip” can be added so that the same check is performed (via *align_epi_anat*.*py*) during the “align” block of processing. The quantitative results of the flip check are automatically parsed and presented to the researcher—along with image snapshots for visual verification—as part of the automatically-generated quality control (QC) HTML output, which *afni_proc*.*py* also creates.

To test the efficacy of this left-right flip test on a range of data, we downloaded publicly available FMRI datasets, both human and non-human (macaque). For variety, datasets were downloaded from a range of locations and projects, with a range of data quality and acquisition parameters; data was only used from subjects who had both an anatomical and FMRI volume in the same session. Human datasets were: OpenFMRI (ds0000003, ds000114, ds000172), FCP (Beijing-Zang, part 1; Cambridge-Buckner, part 1; New York, a/ADHD; Taipei, a) and ABIDE-II (UPSM_Long). Macaque datasets were downloaded from the PRIMate Data Exchange (PRIME-DE; Milham et al., 2018), from the following sites: Institute of Neuroscience (ION), Shanghai; Netherlands Institute of Neuroscience (NIN); Newcastle University; Stem Cell and Brain Research Institute (SBRI); and University of Minnesota (UMinn).

For each study, a subject was randomly assigned to be either “flipped” or “unflipped”, with the anatomical volume left-right flipped for the former group. Each subject’s data were processed using a brief *afni_proc*.*py* command with alignment blocks, including the left-right flip check option (see Appendix A for the full command). The flip results were tallied, and visual verification using the QC HTML output was performed.

## RESULTS

An example^4^ of output from running the left-right flip check as part of FMRI processing with *afni_proc*.*py* is shown in Fig. 2. The figure shows the relevant part of *afni_proc*.*py*’s QC HTML doc, contained within the “warns” block that reports on potential warnings during the processing. The quantitative results are reported (they are also stored in a text or JSON file, for any later use), and in the QC document they are also parsed for immediate identification; also, axial slice montages of each alignment’s result are shown for verification, in the form of the edges of the EPI volume displayed over the original or flipped anatomical. Here, indeed, matching of sulcal and gyral patterns confirm the quantitative flip-check results.

**Figure 2.**
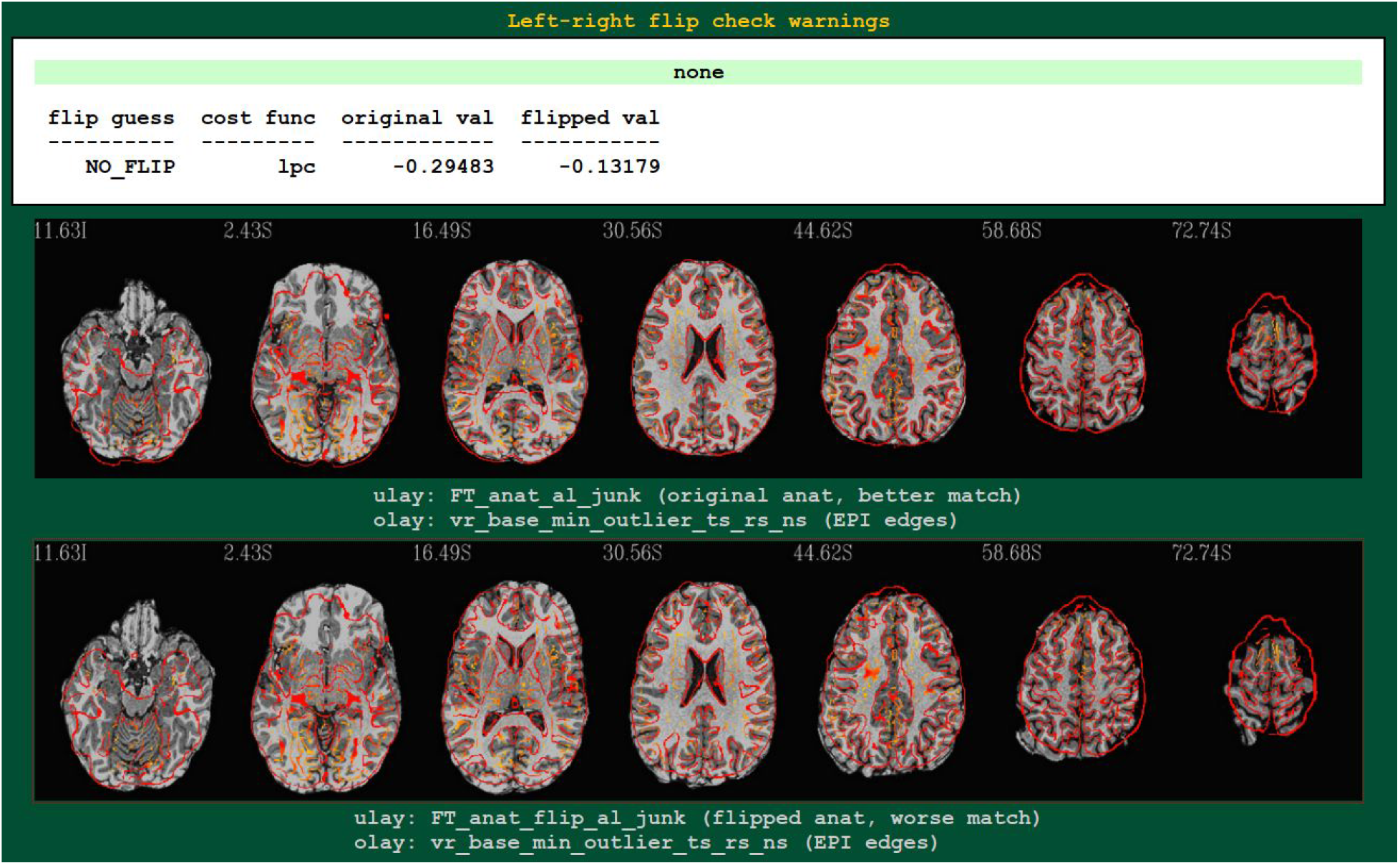
An example of left-right flip checking during FMRI processing with *afni_proc*.*py*, in the automatically generated QC HTML doc. This information is displayed in the “warns” block of the HTML page. The name of the cost function, its values for both cases, and a resulting “guess” evaluation by the program are reported at the top: the “none” in green denotes that NO_FLIP was the result here; a warning in red would be reported there and at the top of the QC page, instead, if DO_FLIP were the result (i.e., if evidence of flipping were found). For visual verification, montages of the results of each alignment are shown as edges of the EPI volume overlaid on each case: original anatomical at top, and flipped version at bottom. In particular, the structures in the superior slices of the cortex provide clear verification of the “flip guess” alignment results.

The results from running the left-right flip check on several publicly available human datasets are summarized in Table 1. Out of 178 subjects analyzed, 100% received the correct left-right flip recommendation: 90/90 subjects in the “flipped” group received DO_FLIP, and 88/88 in the “unflipped” group received NO_FLIP. Visual checks of each subject’s automatically-produced QC HTML (see example in Fig. 2) verified the results.

**Table 1.**
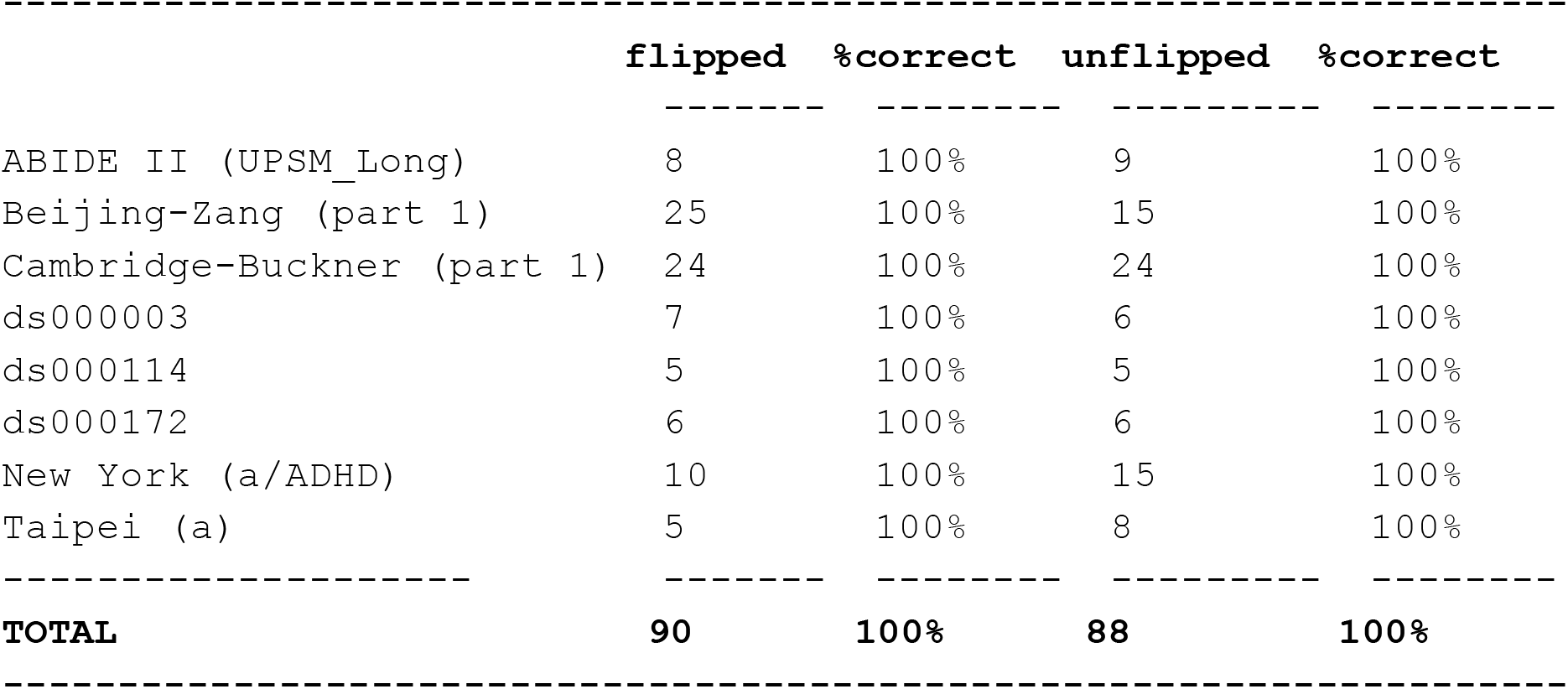
Left-right check (human data). Results of running AFNI’s left-right flip check on several publicly available human datasets using *afni_proc*.*py*. Datasets were randomly assigned to a “flipped” group (anatomical volume left-right flipped before analysis) or to an “unflipped” group (no changes performed). For all datasets, the correct left-right flip results were found; these were verified visually using *afni_proc*.*py*’s automatically generated QC HTML (see Fig. 2).

In the case of macaque datasets, 26 subjects contained both an anatomical and a functional MRI from the same session. Compared to the human datasets, several EPIs had lower tissue contrast, brightness inhomogeneity across the volume and greater distortion (e.g., EPI distortion); additionally, the FOV of EPI and anatomical datasets were shifted in many cases, making alignment much more difficult. In total, either anatomical skullstripping or EPI-anatomical alignment failed in 7 subjects. From the remaining 19 subjects, 7/9 “flipped” and 10/10 “unflipped were correctly categorized (89% correct total); see Table 2. However, it should be noted that the cost function values in 7 of these cases were extremely close and difficult to differentiate. This might be expected due to the aforementioned contrast, brightness and distortion effects, as well as to the inherently high left-right symmetry of anatomy in macaques (much higher than in humans); this symmetry also renders visual verification of the flip check more difficult.

**Table 2.** Left-right check (macaque data). Results of running AFNI’s left-right flip check on several publicly available macaque datasets using *afni_proc*.*py*. Datasets were randomly assigned to a “flipped” group (anatomical volume left-right flipped before analysis) or to an “unflipped” group (no changes performed). While the check correctly categorized datasets in most cases (17 out of 19), the high degree of left-right symmetry in macaques made cost function discrimination difficult for several outputs.

|  | flipped* | %correct | unflipped* | %correct |
| --- | --- | --- | --- | --- |
| ION | 3 | 100% | 2 | 100% |
| Newcastle | 3 | 100% | 6 | 100% |
| NIN | 2 | 50% | 0 | – |
| SBRI | 0 | – | 1 | 100% |
| UMinn | 1 | 0% | 1 | 100% |
| **TOTAL\*\*** | **9** | **78%** | **10** | **100%** |

\* Skullstripping and/or EPI-anatomical alignment failed in 4 additional "flipped" and 3 additional "unflipped" cases (7 out of a total 26 downloaded), and so were not included in this table.
\*\* In 4 "flipped" and 3 "unflipped" cases included in the table (7 out of a total 19 successfully analyzed), the final cost function values were extremely close, making them difficult to differentiate in practice.

In addition to the QC images, one can view the outputs of *align_epi_anat*.*py* (run either directly or via *afni_proc*.*py*) interactively in the AFNI graphical user interface (GUI), and there are several features to aid in the visual comparison of overlaps. While many viewer software packages allow for control of transparency to show one dataset over another, we recommend additional methods for interactive comparisons. Several of these visualization methods are shown in the appendix. Supplementary figure, Fig. A-1 displays several of these for cases of EPI-T1w overlap (differing tissue contrasts) and T2w-DWI comparison (similar tissue contrasts).

## DISCUSSION

Left-right flipping is an unfortunately too common occurrence in MRI analysis. As noted in the Introduction, several of the largest public repositories of publicly available data have had systematically flipped datasets, even after an initial curation process by some of the field’s most experienced researchers. This problem can occur in *any* sized dataset collection, however. In all cases it is incumbent upon the researchers using the data to guard against such problems. We have presented here a simple method for doing so in AFNI, using alignment. This method can be integrated seamlessly into a processing stream (via *afni_proc*.*py*) or checked separately (via *align_epi_anat*.*py*). This approach is efficient, as it takes of order one minute to perform the check. The combination of both quantitative evaluation (using LPC/LPA cost function results) and qualitative verification (using overlaid images of structural features) has performed robustly.

We note that the percent correct (100%) in the tests of public datasets here was higher than expected. Due to the varying quality of tissue contrast, noise and artifacts that can occur in MRI data, we would generally expect some false positives and negatives in the left-right flip checks (which would be verified with visual checks). We also note that no “hidden” flips were found in the data -- i.e., datasets which turned out to have mismatched header information. Based on previous use of the check flip tool introduced here, we had already alerted several repositories, and they have fixed many of the problems. We hope that such curation can continue prospectively in future uploads, and we would recommend the tools presented here for doing so. Such tests are also similarly useful for any center or institute acquiring data, even before it might be analyzed or made public.

It should be noted that this alignment-based method can identify the presence of a *relative* left-right flip between volumes in a dataset. However, it cannot identify an *absolute* left-right flip---that is, it cannot determine which of the volumes has incorrect header information. That kind of information is more difficult to assess, and likely it must be investigated using the original data: checking scan parameters, DICOM conversion and other processing steps. As a corollary, this method cannot detect if *both* volumes in the pair are left-right flipped. However, it may be possible to extend this approach to using an asymmetric MRI template for reference (e.g., the ICBM 2009a/b Nonlinear Asymmetric MNI templates (Fonov et al., 2009)). The degree of variability between subjects and the template would likely reduce the certainty of “absolute flip” detection greatly, particularly across a wide age range or in the case of pathology. Potentially, one could determine left-rightedness based on noted population differences like the petalia, often found in the human brain where the right hemisphere protrudes anteriorly and the left hemisphere protrudes posteriorly (Toga and Thompson, 2003). While it is unlikely that the difference would be suitably reliable on an individual basis, such a biologically-based method could be used as a basis for determining left-right flips over a group of subjects (typically, a group of subjects with the same acquisition and conversion steps will all have the same flip properties).

There can be many causes for left-right flipping (and other header problems) in datasets. As noted above, in cases where flipping *is* detected, a researcher will likely have to backtrack through the provenance and processing of the datasets to find the root cause of the misinformation. For example, DICOM data from scanners can be wrong or ambiguous; the mosaic format of Siemens can have slices stacked in a reversed order, which is in a non-standard part of the file header and hence may not be read properly during conversion. The position of the subject in the scanner must be correctly recorded during acquisition as supine or prone (or “sphinx,” for animal studies). DICOM Conversion PACS systems and custom scripts can also misinterpret information that may have changed in the format, particularly in vendor-specific tags. Older Analyze-7.5 format datasets lack accurate orientation information, and such files may have been used or passed along in studies. NIFTI format datasets have stricter definitions of orientation in their headers, but conflicting sform_matrix and qform_matrix information can still occur, leading to incorrect conversion. Some processing scripts rely on read/write functions in tools that apply either a default orientation or no orientation information, missing consistency checks (e.g., in Matlab, ImageJ or other software); because the NIFTI formats are not part of these codebases’ native format, maintaining correct orientation information in the header is nontrivial and can easily result in mistakes. Some software packages also make assumptions about coordinate storage order and orientation and will either assume the input is in a specific orientation or that multiple input datasets all match each other. Analytical results will have passed through a multitude of steps of conversion, processing and/or regridding in most FMRI and DTI pipelines; maintaining consistency through all steps is a challenge *within* a software package, and combining processing across packages, each with their own assumptions (e.g., some packages ignore such header information), can potentially result in errors.

While the validation of the flip check results of both the human and animal data were consistently high, we note some important differences approaching each of these datasets. Firstly, it should be expected that the flip check results would be much more sensitive in humans, due to the greater left-right asymmetry of structure; indeed, the symmetry of macaque brains made it more difficult to determine consistency with a high degree of confidence (i.e., clearly differentiated cost function and clear visual verification). For any group of subjects with high left-right symmetry, this check becomes less reliable. The quality of data (e.g., amount of artifact, relative tissue contrast, consistent coordinates) also affects the reliability of the check; in the present case, the macaque datasets presented a greater challenge due to such issues. While there are any number of species (human, macaque, rodent, etc.) and types of data (EPI-anatomical, longitudinal anatomical, etc.) to compare, the present results suggest that: for datasets with reasonable quality and left-right asymmetry, this left-right flip check should provide a useful consistency check.

The method presented here provides a simple, fast and verifiable method for an automatic determination of potential left-right flipping problems. In our opinion, it should be included in basic processing streams as a standard feature to detect potential problems in the data. For researchers making data public, it would be particularly beneficial to the community to have run this test. In particular, it may be possible to integrate this with BIDS (Brain Imaging Data Structure) (Gorgolewski et al., 2016) organizational framework and include some basic header validation for coordinate system and orientation.

It should be noted that this left-right flip problem was first observed by an AFNI user (author BRB) while reviewing data, and this highlights an important point: visualizing data remains extremely important in neuroimaging. While we can devise new methods to automatically find some problems (like the left-right flipping issue), there will always be another unforeseen problem that requires carefully looking at the data, and in different ways. We suggest methods that include overlay opacity control, layer toggling, vertical and horizontal curtains, layer blending, checkerboard and edge displays. These kinds of visualization methods help researchers to identify unexpected problems.

The presented method is not foolproof. As noted above, if datasets have poor structural contrast, which may occur fairly often in EPI datasets, for example, then the alignment costs are less reliable. EPI with large flip angles can be a source of the lack of structural information (Gonzalez-Castillo, 2013). Also for data where there is partial coverage, similar lack of structure can occur and potentially decrease the stability of this method.

## CONCLUSIONS

We have found even very basic properties of MRI data like left and right can be confused. Here we presented a simple method to determine consistency among datasets and visualization methods for other unforeseen issues. Even in the era of big data, details still matter—some even more than before, because curating large datasets across multisite studies can pose many new challenges. In the end using a definitive, physical marker while scanning (such as a vitamin E capsule, with a recorded side of placement) is the most robust method to recognize the presence of left-right flipping in a dataset; such a method would also have the benefit of determining flips absolutely. However, to date this practice has not been widely adopted across the neuroimaging community. The presented left-right flip check method in AFNI is simple, efficient and robust. Under the guiding principle of “*caveat emptor*” when using public data, and good practice when using self-acquired data, we strongly recommend the inclusion of this check in all MRI processing pipelines.

## Data Availability

All data is publicly available from open data repositories. The software is open source. Additionally, our processing scripts are provided in the appendix.

https://afni.nimh.nih.gov/pub/dist/edu/data/CD.tgz

https://www.nitrc.org/frs/?group_id=296

https://www.nitrc.org/frs/?group_id=404

https://openneuro.org/datasets/ds000114/versions/1.0.1/download

https://openneuro.org/datasets/ds000172/versions/1.0.1/download

https://openneuro.org/datasets/ds000003/versions/00001/download

http://fcon_1000.projects.nitrc.org/indi/indiPRIME.html

## ACKNOWLEDGEMENTS

We thank O. Esteban and both anonymous reviewers for suggestions to clarify this work. The research and writing of the paper were supported by the NIMH and NINDS Intramural Research Programs (ZICMH002888) of the NIH (HHS, USA), and by the NIH’s Brain Initiative (1R24MH117467-01). This work utilized the computational resources of the NIH HPC Biowulf cluster (http://hpc.nih.gov).

## APPENDIX A Notes on AFNI commands

### Visualization

Visualization methods for comparing two datasets in the AFNI GUI; note that some features are more useful for one case or the other, and several include user interactions in the GUI, as described below). In the first column, one can reduce the opacity of the overlay (olay) volume and investigate where tissue classes appear to overlap (the olay colorbar and range can also be adjusted, as convenient). The second column shows how the underlay (ulay) can be toggled with an “edgified” version of itself using a keypress; this is particularly useful, as often one judges alignment by a comparison of structural boundaries. The third and fourth columns show how one can view both volumes side-by-side, sliding the control bar that appears at the top to control where the boundary (either vertical or horizontal, respectively) appears. In the fifth column the two volumes are blended, with the control bar controlling the relative fraction between 0-100% for the olay. Finally, the sixth column shows how the volumes can be displayed in alternating “checkerboard” squares (this is mostly useful for similar volumes, such as in the lower panel). As each of these modes is turned on/off with a simple key press, one can move among them easily. Methods for generating images automatically as part of an analysis pipeline are noted below in table A-1.

**Figure A-1.**
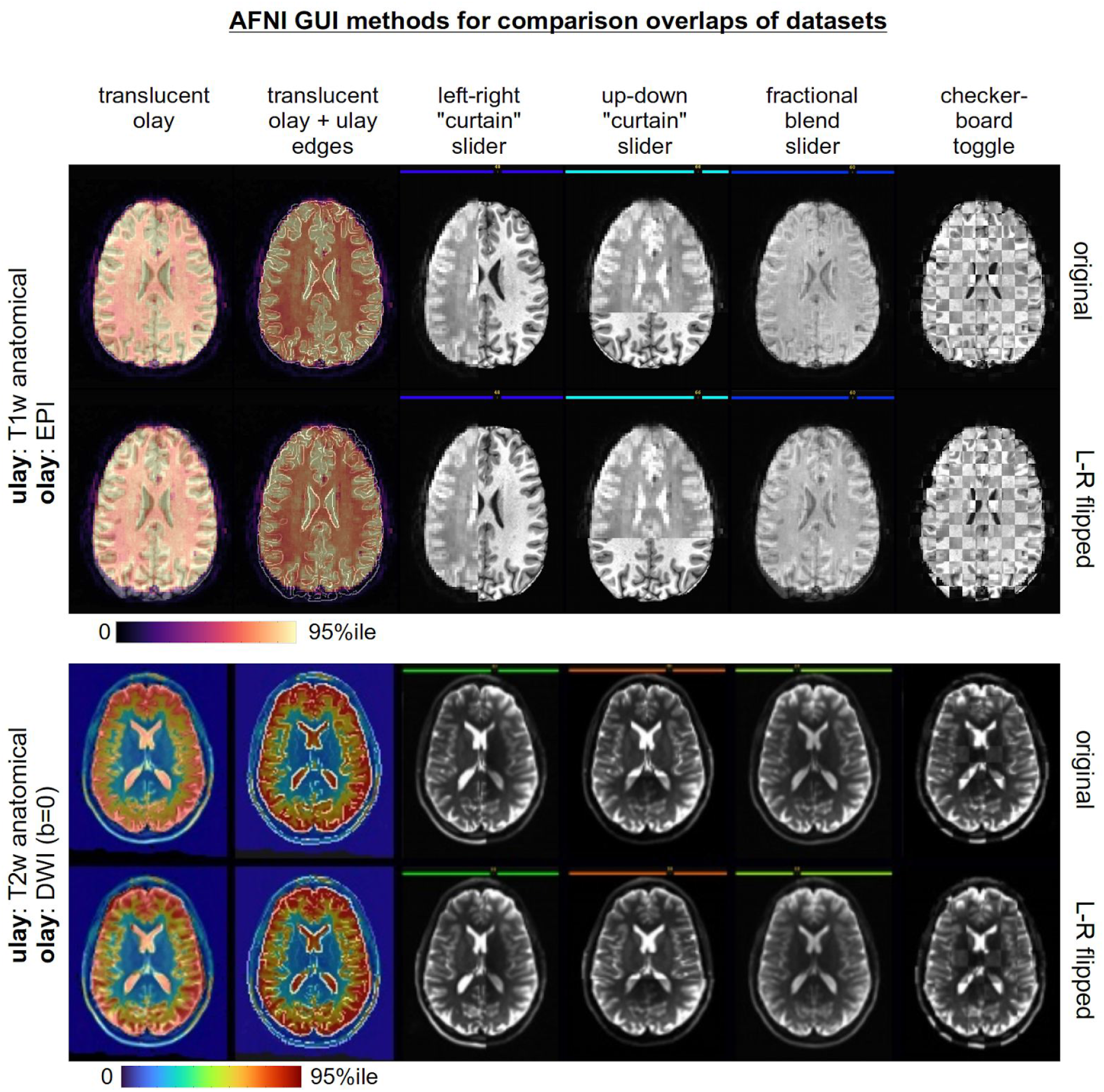
Examples of AFNI GUI features for comparing structural features of two volumes (shown here for axial views, but applicable to all slice planes). Cases where volumes have differing contrasts (T1w-EPI) are compared in the top section, and those with similar contrasts (T2w-DWI) are compared in the lower section. In each case the “original” versions of the volumes appear in the first row, and the versions with a relative left-right flip are shown in the bottom row. In the first column the overlay has opacity set to 4; keypresses are used to enable the features in the remaining columns (from left to right): “e”, “4”, “5”, “6” and “#”. Several of the features allow user interaction in the GUI, such sliding the “curtain” boundary between images or toggling views.

### Processing Pipeline Notes

The *afni_proc*.*py* command used to analyze the public datasets in this study is provided in Table A-1. Some brief comments on the command:

1. The ‘regress’ block is only included to generate the QC, treating the data as resting state. No useful time series analysis is carried out in this particular script.
2. The ‘volreg’ block is mainly included for the ‘MIN_OUTLIER’ functionality: to find and use the EPI volume that has the smallest fraction of outliers for alignment, since it is likely to not be corrupted by subject motion.
3. The ‘-giant_move’ option is included because several datasets had poor initial EPI-anatomical alignment (typically, large relative translations, as well as rotations); this option implements a center of mass alignment to start, and enlarges the parameter search space (3 shifts, 3 angles, 3 scalings, 3 shearings).
4. The lpc+ZZ cost function leads to slightly slower processing than “pure” lpc, but tends to be the most robust for EPI-anatomical alignment.
5. The EPI datasets in the DS000114 group contained two TRs of pre-steady state values, so the value given to the *-tcat_remove_first_trs* option was 2, instead of 0.

**Table A-1.**
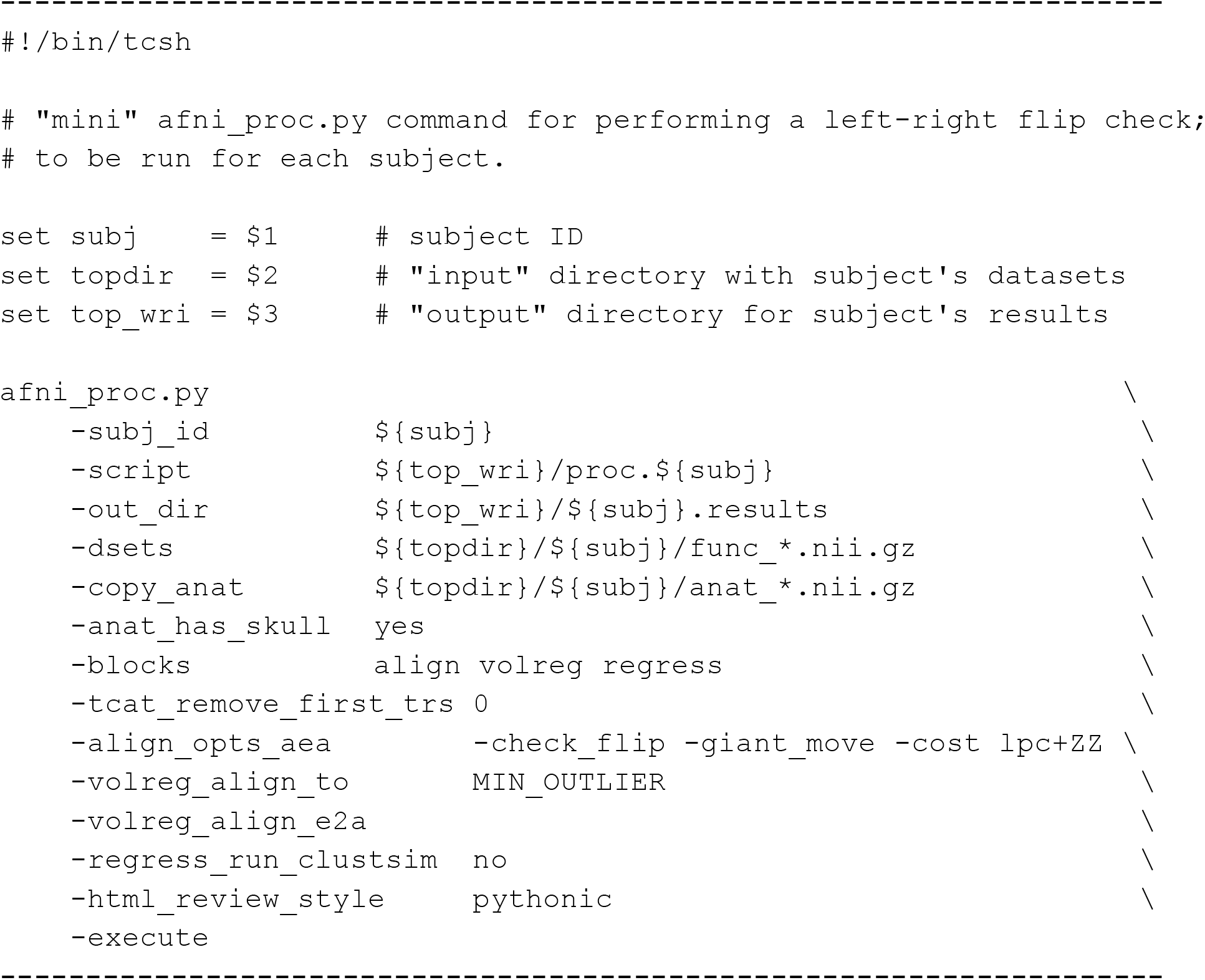
The *afni_proc*.*py* command used to analyze the public data in this study. See comments in the Appendix’s text.

The images in Fig. A-1 can be displayed automatically in the AFNI GUI by creating a “driver” script; this can facilitate the comparison by automating the GUI behavior (loading specific files, jumping to locations, and performing key presses). Additionally, the images in the first two columns can be created and saved without even opening the GUI (e.g., remotely or running on a “headless” system, to be reviewed at any point), through the use of AFNI scripts and commands. In particular, the *@chauffeur_afni* command creates montages across the FOV and can facilitate making systematic sets of views as part of any processing script (for example, it is the tool that creates the images in *afni_proc*.*py’s* HTML QC, shown in part in Fig. 1).

## Footnotes

1 Discussion of orientation issues in FCP datasets: https://www.nitrc.org/forum/forum.php?thread_id=1310&forum_id=1243; Note about dataset orientation fixes in OpenFMRI (after email communication): https://openfmri.org/dataset-orientation-issues/ Note about dataset orientation fixes in ABIDE (after email communication): fcon_1000.projects.nitrc.org/indi/abide/updates/ABIDEII-Usernotes_Updates_Fixes_9_25_16.pdf fcon_1000.projects.nitrc.org/indi/abide/updates/ABIDEII-Usernotes_Updates_Fixes_3_27_17.pdf

2 The initial estimate of LPC+ZZ uses the following formula of cost functions: LPC + 0.4*(HEL + CRA + OV) + 0.2*(NMI + MI), where Hellinger metric, normalized mutual information (NMI), additively symmetrized correlation ratio (CRA), overlap (OV), mutual information (MI) and normalized mutual information (NMI); using multiple cost functions here stabilizes the initial matching. The final stages of alignment are still evaluated using only the LPC cost function, which provides good detail matching due to its designed localness.

3 Its name reflects its original purpose---aligning a subject’s EPI to their T1w anatomical---but it is actually more broadly applicable to any linear affine alignment (typically, to any volumes belonging to the same subject); other programs are recommended for nonlinear alignment.

4 This dataset is publicly available as part of the AFNI Bootcamp demo package (https://afni.nimh.nih.gov/pub/dist/edu/data/CD.tgz), located in the “AFNI_data6/FT_analysis/” directory and processed with afni_proc.py using the accompanying *s05*.*ap*.*uber* script.

## REFERENCES

Bernstein M. (2003). Wrong-side surgery: systems for prevention. Can J Surg.46(2):144–146.

Biswal, et al. (2010). Toward discovery science of human brain function. PNAS 107(10):4734–4739.

Cox RW (1996). AFNI: software for analysis and visualization of functional magnetic resonance neuroimages. Comput. Biomed. Res. 29, 162–173.

Cox RW, Ashburner J, Breman H, Fissell K, Haselgrove C, Holmes CJ, Lancaster JL, Rex DE, Smith SM, Woodward JB, Strother SC (2004). A (sort of) new image data format standard: NIfTI-1. 10th Annual Meeting of the Organization for Human Brain Mapping, 2004.

Digumarthy SR, Vining R, Tabari A, Nandimandalam S, Otrakji A, Shepard JO, Kalra MK (2018). Process improvement for reducing side discrepancies in radiology reports. Acta Radiol Open 7(7-8):2058460118794727. doi: 10.1177/2058460118794727. eCollection 2018 Jul.

DiMartino, et al. (2014). The autism brain imaging data exchange: towards a large-scale evaluation of the intrinsic brain architecture in autism. Mol Psychiatry 19(6):659–67.

Fonov VS, Evans AC, McKinstry RC, Almli CR, Collins DL (2009). Unbiased nonlinear average age-appropriate brain templates from birth to adulthood. NeuroImage 47(1):S102.

Gonzalez-Castillo, J., Duthie, K. N., Saad, Z. S., Chu, C., Bandettini, P. A., & Luh, W. M. (2013). Effects of image contrast on functional MRI image registration. NeuroImage, 67, 163–174. doi:10.1016/j.neuroimage.2012.10.076

Gorgolewski KJ, et al. (2016). The brain imaging data structure, a format for organizing and describing outputs of neuroimaging experiments. Sci Data 3, 160044.

Landau E, Hirschorn D, Koutras I, Malek A, Demissie S (2015). Preventing errors in laterality. J Digit Imaging. 28(2):240–246. doi:10.1007/s10278-014-9738-4

Milham MP, et al. An open resource for non-human primate imaging. Neuron. 2018;100:61–74.e2. doi: 10.1016/j.neuron.2018.08.039.

Poldrack RA, et al. (2013). Toward open sharing of task-based fMRI data: the OpenfMRI project. Frontiers in Neuroinformatics 7:12. doi:10.3389/fninf.2013.00012.

Saad ZS, Glen DR, Chen G, Beauchamp MS, Desai R, Cox RW (2009). A new method for improving functional-to-structural MRI alignment using local Pearson correlation. Neuroimage 44(3):839–48.

Sangwaiya MJ, Saini S, Blake MA, Dreyer KJ, Kalra MK (2009). Errare humanum est: frequency of laterality errors in radiology reports. AJR Am J Roentgenol 192(5):W239–44.

Toga AW, Thompson PM, (2003). Mapping brain asymmetry. Nature Reviews Neuroscience 4,37–48.

